# Are female-specific cancers long-term sequelae of COVID-19? Evidence from a large-scale genome-wide cross-trait analysis

**DOI:** 10.1101/2022.08.25.22279195

**Authors:** Xunying Zhao, Xueyao Wu, Jinyu Xiao, Li Zhang, Yu Hao, Chenghan Xiao, Ben Zhang, Jiayuan Li, Xia Jiang

**Author notes:** **Correspondence to:** West China School of Public Health and West China Fourth Hospital, Sichuan University 16#, Section 3, Renmin Nan Lu Chengdu, Sichuan 610041, P. R. China (Ben Zhang, PhD,; Jiayuan Li, PhD,; Xia Jiang, PhD,). Xunying Zhao and Xueyao Wu contributed equally to this work.

## Abstract

**Background:** Little is known regarding the long-term adverse effects of COVID-19 on female-specific cancers due to the restricted length of observational time, nor the shared genetic influences underlying these conditions.

**Methods:** Leveraging summary statistics from the hitherto largest genome-wide association studies conducted in each trait, we performed a comprehensive genome-wide cross-trait analysis to investigate the shared genetic architecture and the putative genetic associations between COVID-19 with three main female-specific cancers: breast cancer (BC), epithelial ovarian cancer (EOC), and endometrial cancer (EC). Three phenotypes were selected to represent COVID-19 susceptibility (SARS-CoV-2 infection) and severity (COVID-19 hospitalization, COVID-19 critical illness).

**Results:** For COVID-19 susceptibility, we found no evidence of a genetic correlation with any of the female-specific cancers. For COVID-19 severity, we identified a significant genome-wide genetic correlation with EC for both hospitalization (*r*_*g*_=0.19, *P*=0.01) and critical illness (*r*_*g*_ =0.29, *P*=3.00×10^−4^). Mendelian randomization demonstrated no valid association of COVID-19 with any cancer of interest, except for suggestive associations of genetically predicted hospitalization (OR_IVW_=1.09, 95%CI=1.01-1.18, *P*=0.04) and critical illness (OR_IVW_=1.06, 95%CI=1.00-1.11, *P*=0.04) with EC risk, none withstanding multiple correction. No reverse association was found. Cross-trait meta-analysis identified multiple pleiotropic SNPs between COVID-19 and female-specific cancers, including 20 for BC, 15 for EOC, and 5 for EC. Transcriptome-wide association studies revealed shared genes, mostly enriched in the hematologic, cardiovascular, and nervous systems.

**Conclusions:** Our genetic analysis highlights an intrinsic link underlying female-specific cancers and COVID-19 - while COVID-19 is not likely to elevate the immediate risk of the examined female-specific cancers, it appears to share mechanistic pathways with these conditions. These findings may provide implications for future therapeutic strategies and public health actions.

## Introduction

With more than half a billion registered infections and 6.4 million deaths globally, the coronavirus disease 2019 (COVID-19) continues to spread (WHO: https://covid19.who.int/). Despite lungs being the organ predominately affected, a multi-system involvement of COVID-19 is well-characterized, with extra-pneumatic manifestations documented in hematologic, cardiovascular, neurological tissues, and others, possibly caused by direct viral virulence or as a result of immunopathological reactions^1,2^. Moreover, while most COVID-19 patients recover within a couple of weeks after infection, a non-negligible proportion of individuals experience chronic symptoms lasting for months, especially in women^3,4^. Given the extensive and likely prolonged impairment of COVID-19 on multiple bodily systems, its long-term sequelae have become increasingly recognized and concerning^3,5^.

Strong evidence has been raised reflecting the disparities in COVID-19 pandemic, potentially mediated through unique social determinants of health^6,7^. Women, especially those with high health burdens are affected disproportionally by COVID-19^6,8^. For example, individuals with breast cancer (BC) were nearly 3-times more likely to die from COVID-19 than their non-cancer referents (odds ratio, OR = 3.30; 95% confidence interval, 95%CI = 1.96-5.57)^9^. Among women with gynecological cancers, mainly epithelial ovarian cancer (EOC) and endometrial cancer (EC), a significantly increased mortality due to COVID-19 (14.0%)^10^ was found compared to general population (5.6%)^11^. Indeed, several shared signaling pathways, including cytokine, immunosuppression, coagulation disorders, inflammatory reactions, and hormone secretion^12-15^, have been reported. Nevertheless, whether COVID-19 increases the susceptibility to cancer in those without prior malignancies remains unclear due to the hitherto restricted length of observational time. It is concerned that COVID-19 may predispose recovered patients to cancer development based on the growing evidence of SARS-CoV-2 in modulating oncogenic pathways, promoting chronic low-grade inflammation, and causing tissue damage^12,16^.

One way of evaluating the putative causal association underlying two phenotypes is to apply Mendelian randomization (MR)^17^, a framework leveraging genetic variants as instruments to overcome the limitation of conventional epidemiological designs, such as restricted observational duration, environmental confounder, and reverse association^18^. Using other genetic methods including genetic correlation analysis^19^, cross-trait meta-analysis^20^, and transcriptome-wide association study (TWAS)^21^, shared genetic influences across traits can also be quantified, driving forward epidemiologic associations with novel insights into the underlying biological mechanisms. Here, we apply these methods to perform a comprehensive genome-wide cross-trait analysis^18^, with an overarching goal of characterizing the shared genetic architecture and the putative associations underpinning COVID-19 and female-specific cancers (BC, EOC, and EC). Three COVID-19 phenotypes were included, namely SARS-CoV-2 infection, COVID-19 hospitalization, and COVID-19 critical illness. The overview of study design is shown in **Fig 1**.

**Fig 1.**
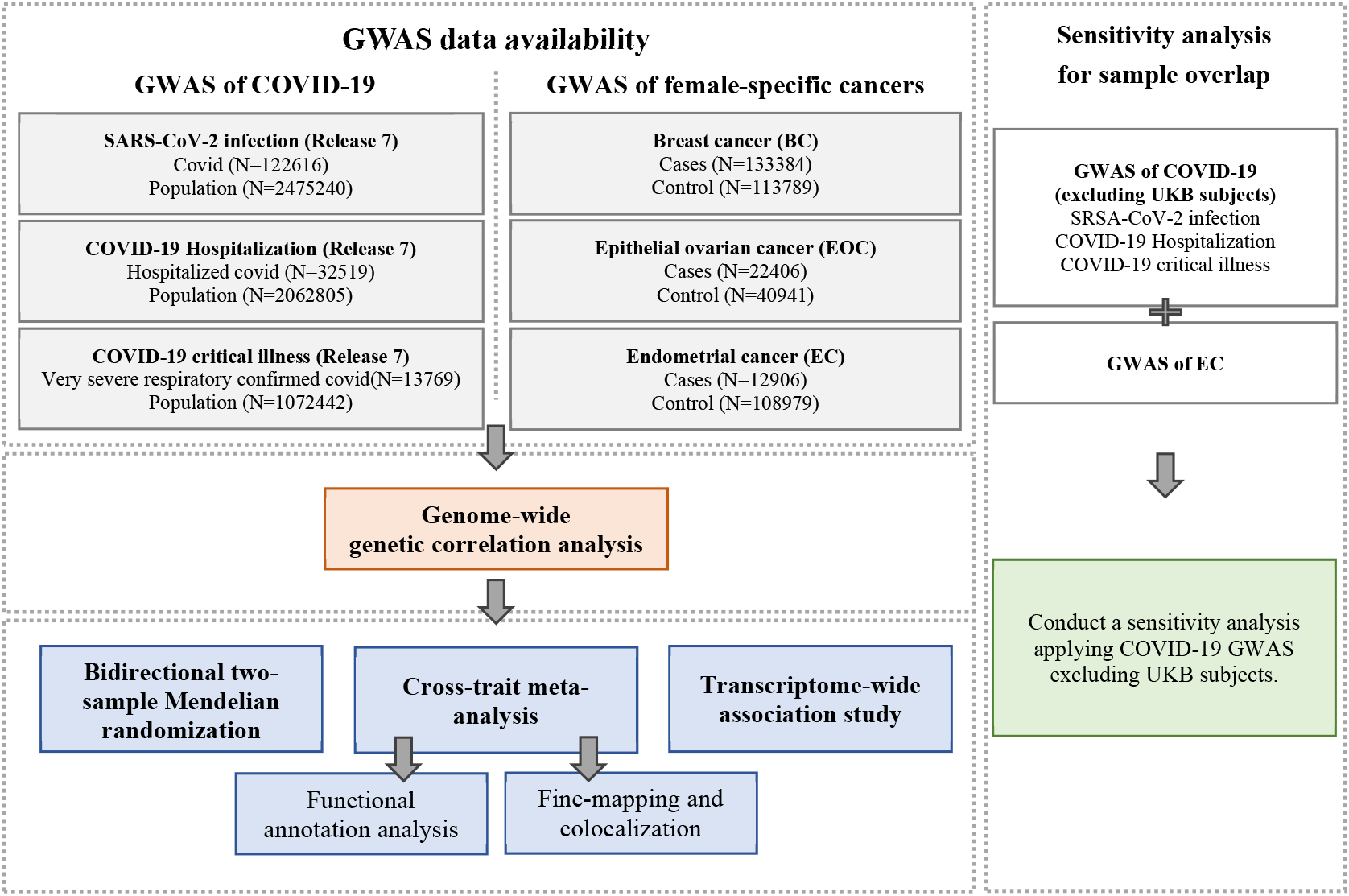
Overall study design of genome-wide cross-trait analysis. GWAS summary statistics for each trait of interest were retrieved from publicly available GWAS(s). GWAS: genome-wide association study; UKB: UK Biobank.

## Materials and methods

### GWAS data sets

#### Cancer

Three common female malignant tumors, the breast cancer (BC), the epithelial ovarian cancer (EOC), and the endometrial cancer (EC)^22^, were included in our study. GWAS summary data of BC was obtained from a meta-analysis of the Breast Cancer Association Consortium (BCAC) and 11 other BC genetic studies^23^, involving 133,384 cases and 113,789 controls. GWAS summary data of overall invasive EOC was obtained from the Ovarian Cancer Association Consortium (OCAC) meta-analysis^24^, involving 22,406 cases and 40,941 controls. GWAS summary data of EC was obtained from a meta-analysis of the Endometrial Cancer Association Consortium (ECAC), the Epidemiology of Endometrial Cancer Consortium (E2C2), and the UK Biobank^25^, involving 12,906 cases and 108,979 controls. All individuals were of European ancestry.

#### COVID-19

For COVID-19 phenotypes, we used the hitherto largest GWAS summary data of European ancestry conducted by the COVID-19 Host Genetics Initiative, release 7 (https://www.covid19hg.org/), from which subjects of 23andMe were excluded due to data restrictions^26^. Three phenotypes were selected and further divided into two categories, representing COVID-19 susceptibility and severity. **SARS-CoV-2 infection**, defined as “cases with reported SARS-CoV-2 infection regardless of symptoms (N = 122,616) vs. population (N = 2,475,240)”, was used to index COVID-19 susceptibility. **COVID-19 hospitalization**, defined as “moderate or severe COVID-19 patients who were hospitalized due to COVID-19 symptoms (N = 32,519) vs. population (N = 2,062,805)”, and **COVID-19 critical illness**, defined as “severe COVID-19 patients who needed respiratory support or who died due to the disease (N = 13,769) vs. population (N = 1,072,442)” were used to index COVID-19 severity. Considering a potential sample overlap between GWAS of EC and GWAS of COVID-19 phenotypes (both involving UK Biobank subjects), we further performed a sensitivity analysis using trans-ancestry COVID-19 GWAS excluding individuals of UKB. Details on the characteristics of each included dataset are presented in **Supplementary Table 1**.

### Statistical analysis

#### Genome-wide genetic correlation analysis

To describe the average shared genetic effect between female-specific cancers and COVID-19 phenotypes, we quantified their genome-wide genetic correlation using pairwise linkage-disequilibrium score regression (LDSC)^19^. The genetic correlation estimates *r*_*g*_ range from -1 to +1, with +1 indicating a complete positive correlation and -1 indicating a complete negative correlation. We used pre-computed LD-scores obtained from ∼1.2 million common SNPs of European ancestry represented in the Hapmap3 reference panel. Bonferroni correction was applied to account for multiple testing. We defined a significant *r*_*g*_ as *P* < 5.56×10^−3^ (α = 0.05/9, number of phenotype pairs)^27^, and suggestive *r*_*g*_ as 5.56×10^−3^ ≤ *P* < 0.05.

#### Bidirectional Mendelian randomization analysis

The average shared genetic effects can be decomposed into vertical pleiotropy and/or horizontal pleiotropy, where vertical pleiotropy (or a putative causal association) refers to genetic variants affecting one trait (outcome) via its effect on an intermediate trait (exposure), and horizontal pleiotropy, often simplified as pleiotropy, refers to genetic variants affecting both traits independently^18^. To further explore these alternatives, we first conducted a bidirectional two-sample MR between COVID-19 phenotypes and female-specific cancers. As no significant SNP was reported in the original COVID-19 GWAS, we selected independent instrumental variables (IVs) by clumping all variants that reached genome-wide significance (*P* < 5×10^−8^) according to a strict criterion (*r*^2^ ≤ 0.001 within a 1.0Mb window). For cancers, we collected all previously reported independent index SNPs reaching genome-wide significance (*P* < 5×10^−8^) from corresponding GWAS. We calculated the *F*-statistic to evaluate instrument strength, with an *F*-statistic < 10 indicating a weak instrument^28^.

We applied inverse-variance weighted (IVW) method as our primary approach^17^, complemented with MR-Egger^29^ and weighted median^30^ to evaluate its robustness. A Bonferroni-corrected significance threshold of *P* < 5.56×10^−3^ (α = 0.05/9, number of phenotype pairs) was applied^27^, while 5.56×10^−3^ ≤ *P* < 0.05 was defined as suggestive significance. An MR effect estimate was considered robust if it was statistically significant in IVW and remained directionally consistent across both the MR-Egger and the weighted median approaches.

To validate MR model assumptions, we conducted several important sensitivity analyses. First, we excluded palindromic IVs that have the same alleles on forward and reverse strands, and pleiotropic IVs that are associated with potential confounders according to GWAS catalog (https://www.ebi.ac.uk/gwas/, accessed on 05/08/2021). Next, we performed a leave-one-out analysis in which we excluded one IV at a time and conducted IVW using the remaining SNPs to identify outlying instruments. Finally, we used MR-Pleiotropy Residual Sum and Outlier (MR-PRESSO) approach to detect and correct for horizontal pleiotropy^31^.

#### Cross-trait meta-analysis

To identify pleiotropic loci affecting both traits, we further performed a cross-trait meta-analysis using Cross Phenotype Association (CPASSOC)^20^. We chose S_Het_, a statistic that is more powerful for heterogonous effects (common when meta-analysing different traits), to combine summary statistics across traits. We used PLINK 1.9 “clumping” function to obtain independent loci with parameters: --clump-p1 5e-8 --clump-p2 1e-5 --clump-r2 0.2 --clump-kb 500^32^. Index SNPs, satisfying *P*_CAPSSOC_ < 5×10^−8^ and *P*_single-trait_ < 1×10^−3^ (both traits), were considered as significant pleiotropic SNPs. An index SNP satisfying the following conditions was considered as a novel shared SNP: (1) did not reach genome-wide significance (5×10^−8^ < *P*_single-trait_ < 1×10^−3^) in single-trait GWAS; and (2) was not in LD (*r*^2^ < 0.05) with any of the previously reported genome-wide significant SNPs in single-trait GWAS. To further investigate biological insights for the shared variants, we use Ensemble Variant Effect Predictor (VEP) to annotate the linear closest genes of the identified pleiotropic SNPs^33^.

#### Fine-mapping credible set and colocalization analysis

Due to the complex LD patterns among SNPs, index SNPs are not necessarily causal variations^34^. We conducted a fine-mapping analysis using FM-summary to identify a credible set of variants that were 99% likely to contain causal variants at each of the shared loci. FM-summary is a fine-mapping algorithm in Bayesian framework which maps the primary signal and uses a flat prior with steepest descent approximation^35^.

To assess whether the same variants are responsible for two GWAS signals or are distinct variants close to each other, we conducted a colocalization analysis using Coloc^36^. Coloc is a tool in Bayesian framework that provides intuitive posterior probabilities of 5 hypotheses (H0-H4). We extracted summary statistics for variants within 500 kb of each shared index SNP and calculated the posterior probability for H4 (PPH4, the probability that both traits are associated through sharing a single causal variant). A locus was considered colocalized if PPH4 was greater than 0.5.

#### Transcriptome-wide association studies

Many genetic variants implement their function by modulating gene expression in different tissues, thus, considering gene expression and tissue specificity help clarify common biological mechanisms. We performed TWAS to identify genes whose expression is significantly associated with traits using FUSION^21^. We integrated GWAS summary data with expression weights across 48 tissues from GTEx (Genotype-Tissue Expression, version 7), with one tissue-trait pair at a time. Bonferroni-correction was applied for all gene-tissue pairs tested (∼230,000 in total) within each trait, and *P*_Bonferroni_ < 0.05 was defined as significance^27^. To identify an independent set of gene-tissue pairs, we conducted joint/conditional tests (an extension of TWAS) among regions with multiple identified signals^37^. Shared gene-tissue pairs were determined through intersection across traits.

## Results

### Genome-wide genetic correlation

For COVID-19 susceptibility, we found no evidence on a shared genetic basis with any of the female-specific cancers (BC: *r*_*g*_ = *-*0.01, *P* = 0.90; EOC: *r*_*g*_ = 0.01, *P* = 0.91; EC: *r*_*g*_ = 0.09, *P* = 0.23; **Table 1**). For COVID-19 severity, we identified a suggestive genetic correlation for hospitalization with EC (*r*_*g*_ = 0.19, *P* = 0.01), as well as a significant genetic correlation for critical illness with EC (*r*_*g*_ = 0.29, *P* = 3.00×10^−4^). No significant result was found for COVID-19 severity with either BC (hospitalization: *r*_*g*_ = 0.06, *P* = 0.16; critical illness: *r*_*g*_ = 0.05, *P* = 0.28) or EOC (hospitalization: *r*_*g*_ = *-*0.04, *P* = 0.55; critical illness: *r*_*g*_ = *-*0.02, *P* = 0.77). Interestingly, for EC and COVID-19, both the magnitude and the significance of *r*_*g*_ increased as the disease developed, from infection (0.09) to hospitalization (0.19) to critical illness (0.29).

**Table 1.** Genetic correlation between female-specific cancers and COVID-19 phenotypes.

| Cancer | COVID-19 phenotype | $r_g$ | 95%CI | P-value |
| --- | --- | --- | --- | --- |
| BC | Infection | -0.01 | (-0.09,0.08) | 0.90 |
|  | Hospitalization | 0.06 | (-0.02,0.14) | 0.16 |
|  | Critical illness | 0.05 | (-0.04,0.13) | 0.28 |
| EOC | Infection | 0.01 | (-0.16,0.18) | 0.91 |
|  | Hospitalization | -0.04 | (-0.19,0.10) | 0.55 |
|  | Critical illness | -0.02 | (-0.17,0.13) | 0.77 |
| EC | Infection | 0.09 | (-0.06,0.24) | 0.23 |
|  | **Hospitalization** | **0.19** | **(0.04,0.34)** | **0.01** |
|  | **Critical illness** | **0.29** | **(0.14,0.45)** | **$3.00 \times 10^{-4*}$** |

Bold-face: $P < 0.05$ ; $*P < 5.56 \times 10^{-3}$ .
$r_g$ : genetic correlation; CI: confidence interval; Infection: reported SARS-CoV-2 infection vs. population; Hospitalization: COVID-19 hospitalization patients vs. population; Critical illness: very severe respiratory confirmed COVID-19 patients vs. population; BC: breast cancer; EOC: overall invasive epithelial ovarian cancer; EC: endometrial cancer

### Bidirectional Mendelian randomization

We continued to conduct a MR to evaluate potential associations of genetically predicted COVID-19 phenotypes on female-specific cancers risk, motivated by the significant shared genetic basis. We identified and selected 16, 38, and 37 SNPs as IVs for infection, hospitalization, and critical illness of COVID-19. *F*-statistics suggested minimal weak instrument bias (**Supplementary Table 2**). ***For COVID-19 susceptibility***, we did not find any association with female-specific cancer (BC: OR_IVW_ = 0.99, 95%CI = 0.86-1.14, *P* = 0.92; EOC: OR_IVW_ = 1.19, 95%CI = 0.95-1.48, *P* = 0.12; EC: OR_IVW_ = 1.26, 95%CI = 0.97-1.64, *P* = 0.09; **Fig 2** and **Supplementary Table 3-4**). ***For COVID-19 severity***, genetically predicted hospitalization (OR_IVW_ = 1.09, 95%CI = 1.01-1.18, *P* = 0.04) and critical illness (OR_IVW_ = 1.06, 95%CI = 1.00-1.11, *P* = 0.04) were associated with the risk of EC under suggestive significance, none of which withstood multiple correction. The estimates remained directionally consistent using the MR-Egger regression (hospitalization: OR = 1.06, 95%CI = 0.91-1.24; critical illness: OR = 1.03, 95%CI = 0.93-1.13) or the weighted median approach (hospitalization: OR = 1.05, 95%CI = 0.95-1.16; critical illness: OR = 1.03, 95%CI = 0.97-1.10). No substantial alteration was found after excluding palindromic SNPs or pleiotropic SNPs, and the leave-one-out analysis demonstrated that the pooled estimate was not driven by any outlying variant. MR-PRESSO yielded to similar findings. No association of genetically predicted COVID-19 severity was found for BC (hospitalization: OR_IVW_ = 1.00, 95%CI = 0.96-1.05; critical illness: OR_IVW_ = 1.01, 95%CI = 0.98-1.04) or EOC (hospitalization: OR_IVW_ = 1.01, 95%CI = 0.92-1.10; critical illness: OR_IVW_ = 0.99, 95%CI = 0.92-1.06).

**Fig 2.**
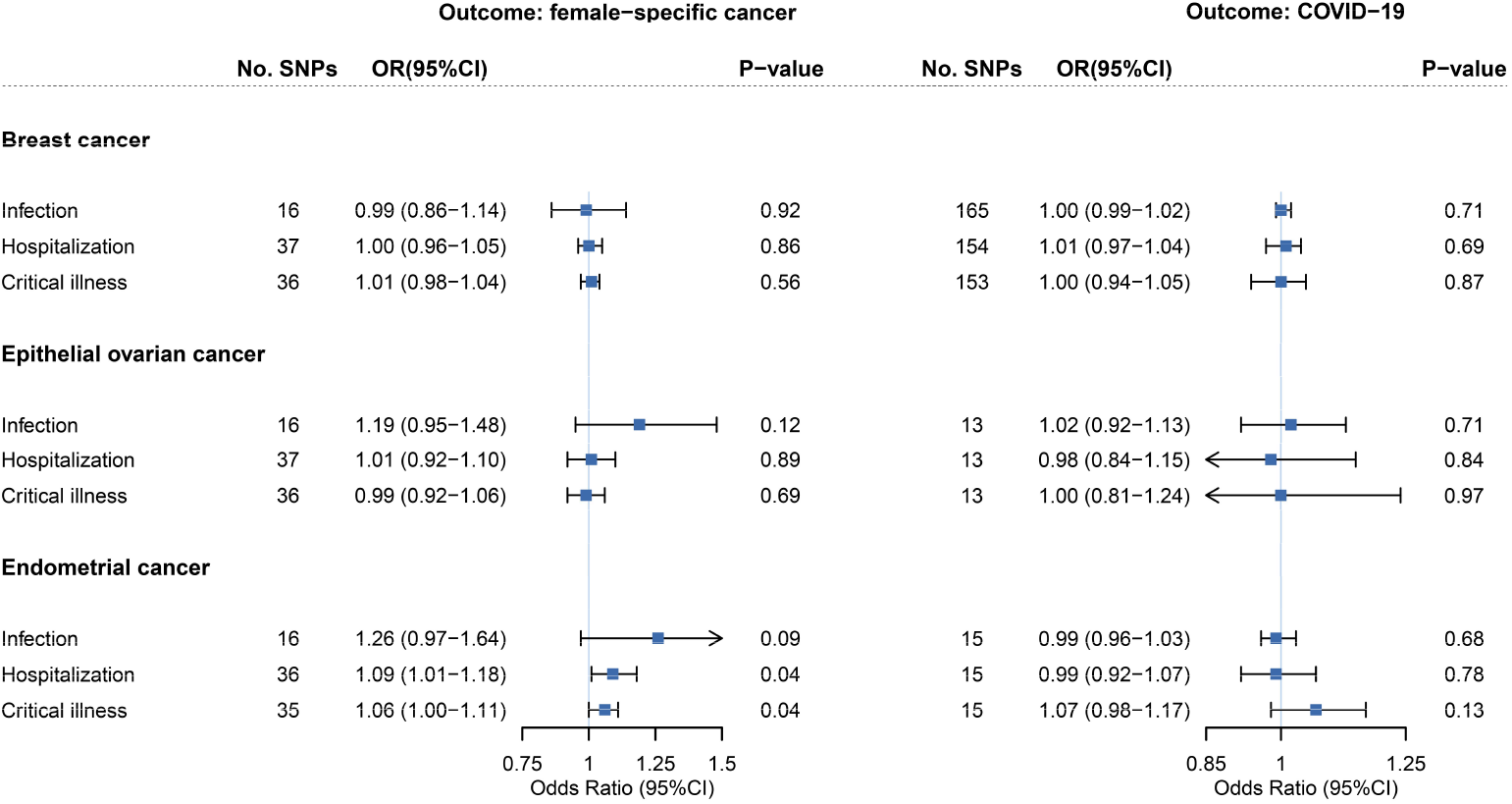
Bidirectional Mendelian randomization associations between COVID-19 phenotypes and female-specific cancers. On the left are the MR effect estimates of genetically predicted COVID-19 phenotypes on each female-specific cancer by the inverse-variance weighted approach. On the right are the MR effect estimates of genetically predicted female-specific cancer on COVID-19 phenotypes by the inverse-variance weighted approach. Boxes represent the point estimates of MR effects, and error bars represent 95% confidence intervals.

In the reverse-direction MR where female-specific cancers were considered as exposures, we selected 168, 13, and 16 SNPs as IVs to proxy BC, EOC, and EC. *F*-statistics for these IVs suggested strong instruments (**Supplementary Table 5**). None of the three genetically predicted female-specific cancers appeared to affect COVID-19 susceptibility or severity (**Fig 2** and **Supplementary Table 6**).

### Cross-trait meta-analysis and pleiotropic loci

With little sign of vertical pleiotropy, we continued to perform cross-trait meta-analysis to reveal horizontal pleiotropic effect of individual variant. A total of 20 independent pleiotropic SNPs were identified as shared by BC with at least one COVID-19 phenotype, including 7 for infection, 7 for hospitalization, and 6 for critical illness (**Fig 3** and **Supplementary Table 7-10**). These 20 SNPs were mainly located at genomic regions 17q21.31 (harboring *WNT3, MAPT, CRHR1*, and *PLEKHM1*), 9q34.2 (harboring *ABO, LCN1P2*, and *REXO4*), and 1q22 (harboring *THBS3, GON4L*, and *PMF1*). Notably, SNP rs910416 located at 6q25.1 showed the most significance (*P*_CPASSOC_ = 1.90×10^−29^), followed by SNP rs17474001 at 9q34.2 (*P*_CPASSOC_ = 2.48×10^−26^), and SNP rs9411395 at 9q34.2 (*P*_CPASSOC_ = 4.71×10^−26^). We also identified a novel shared locus (index SNP rs1052067, *P*_CPASSOC_ = 2.76×10^−08^) located at 1q22.

**Fig 3.**
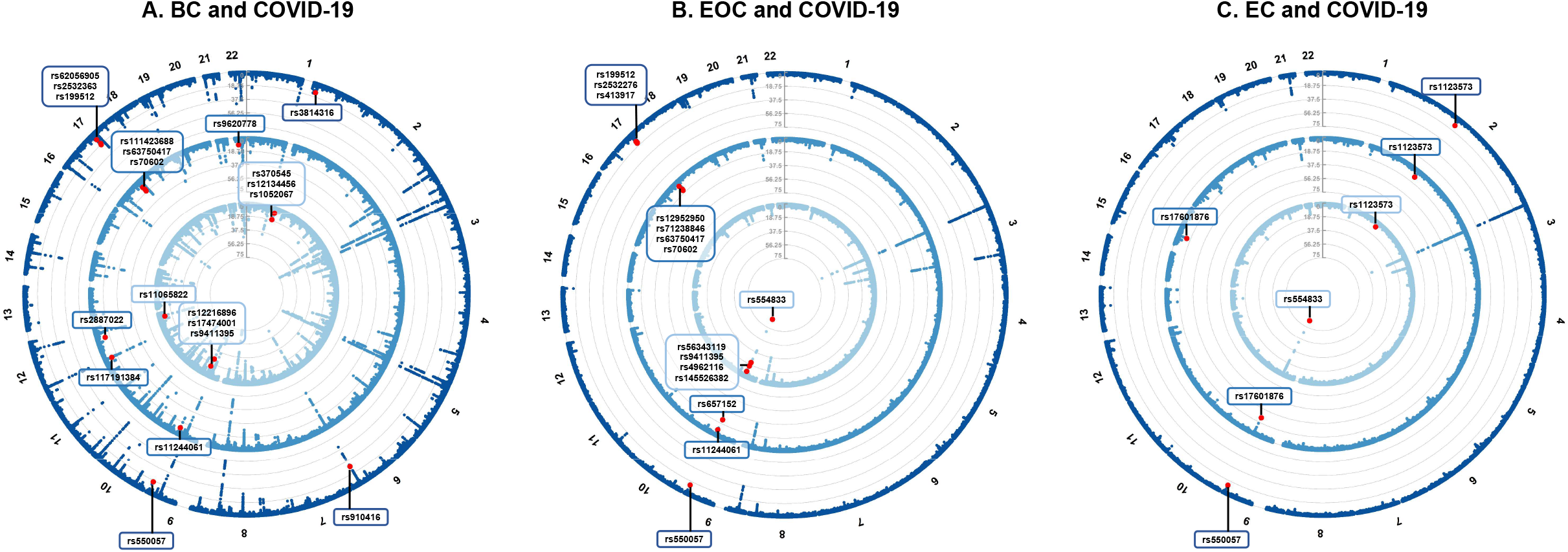
Pleiotropic loci between female-specific cancers and COVID-19 phenotypes identified from cross-trait meta-analysis. (A) Pleiotropic loci identified for breast cancer and COVID-19 phenotypes; (B) pleiotropic loci identified for epithelial ovarian cancer and COVID-19 phenotypes; (C) pleiotropic loci identified for endometrial cancer and COVID-19 phenotypes. In each circular Manhattan plot, the circle from center to periphery shows the cross-trait meta-analysis results between each female-specific cancer and the three COVID-19 phenotypes (light blue: SRAS-CoV-2 infection, blue: COVID-19 hospitalization, dark blue: COVID-19 critical illness). The outermost numbers represent chromosomes 1-22. The red dots represent significant pleiotropic SNPs in cross-trait meta-analysis (*P*_CPASSOC_ < 5×10^−8^ and *P*_single-trait_<1×10^−3^ in both traits).

A total of 15 independent pleiotropic SNPs were identified as shared by EOC with at least one COVID-19 phenotype, including 5 for infection, 6 for hospitalization, and 4 for critical illness. These 15 SNPs were mainly distributed at 2 loci, 9q34.2 (harboring *ABO, SURF4*, and *LCN1P2*) and 17q21.31 (harboring *WNT3, MAPT, CRHR1*, and *PLEKHM1*). All top-three most significant SNPs were located at 9q34.2, including SNP rs554833 (*P*_CPASSOC_ = 1.28×10^−88^), SNP rs657152 (*P*_CPASSOC_ = 6.25×10^−25^), and SNP rs56343119 (*P*_CPASSOC_ = 1.28×10^−22^).

A total of 5 independent pleiotropic SNPs were identified as shared by EC with at least one COVID-19 phenotype, including 2 for infection, 3 for hospitalization, and 2 for critical illness. Three out of the 5 shared SNPs were located at 9q34.2 (harboring *ABO*). Among the rest, SNP rs1123573 was located at 2p16.1 (harboring *BCL11A*), and SNP rs17601876 was located at 15q21.2 (harboring *CYP19A1, RP11-108K3*.*1*). Index SNP rs554833 at 9q34.2 showed the most significance (*P*_CPASSOC_ = 3.29×10^−85^), followed by SNP rs657152 at 9q34.2 (*P*_CPASSOC_ = 3.77×10^−26^), and SNP rs17601876 at 15q21.2 (*P*_CPASSOC_ = 3.07×10^−14^).

### Identification of causal variants and colocalization

For all identified pleiotropic SNPs, we determined a 99% credible set of causal SNPs using FM-summary. A total of 4568 candidate SNPs were identified as the credible set of shared causal SNPs for BC and COVID-19 phenotypes. Corresponding figures in EOC and EC were 4893 and 106. Particularly, we identified only one candidate causal SNP in the 99% credible set for BC/infection (rs12216896), BC/hospitalization (rs2887022), and EC/hospitalization (rs1123573). Lists of candidate causal SNPs at each pleiotropic locus were shown in **Supplementary Table 11-13**.

We next performed colocalization analysis to determine whether genetic variants driving the association between different traits are the same. We identified several loci to colocalize at the same candidate SNPs (PPH4 > 0.5), including 4 shared loci for BC and COVID-19 phenotypes, 8 shared loci for EOC and COVID-19 phenotypes, and 3 shared loci for EC and COVID-19 phenotypes (**Supplementary Table 14**).

### Transcriptome-wide association studies and shared genes

We identified multiple independent gene-tissue pairs shared between female-specific cancers and COVID-19 phenotypes (**Table 2**).

**Table 2.**
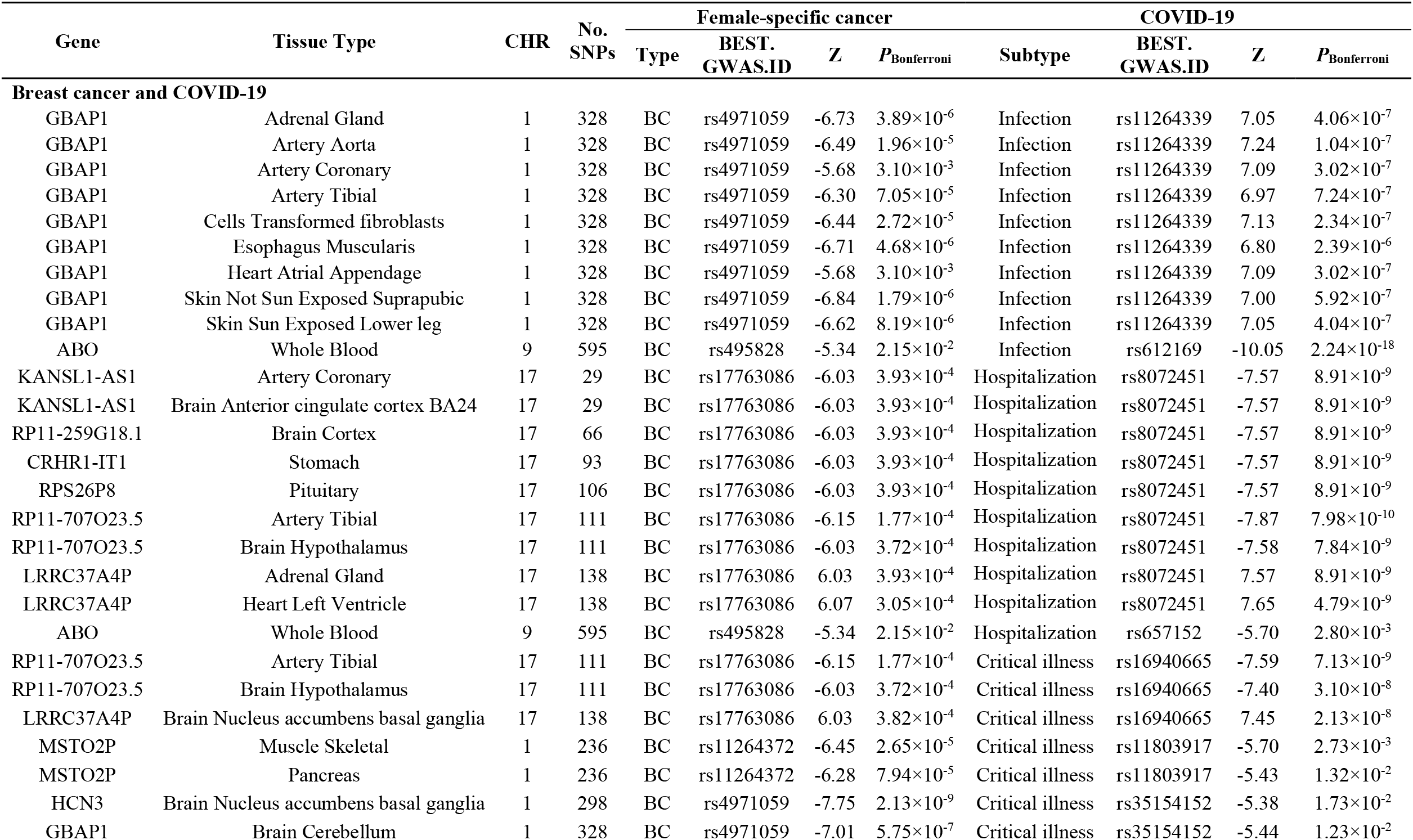
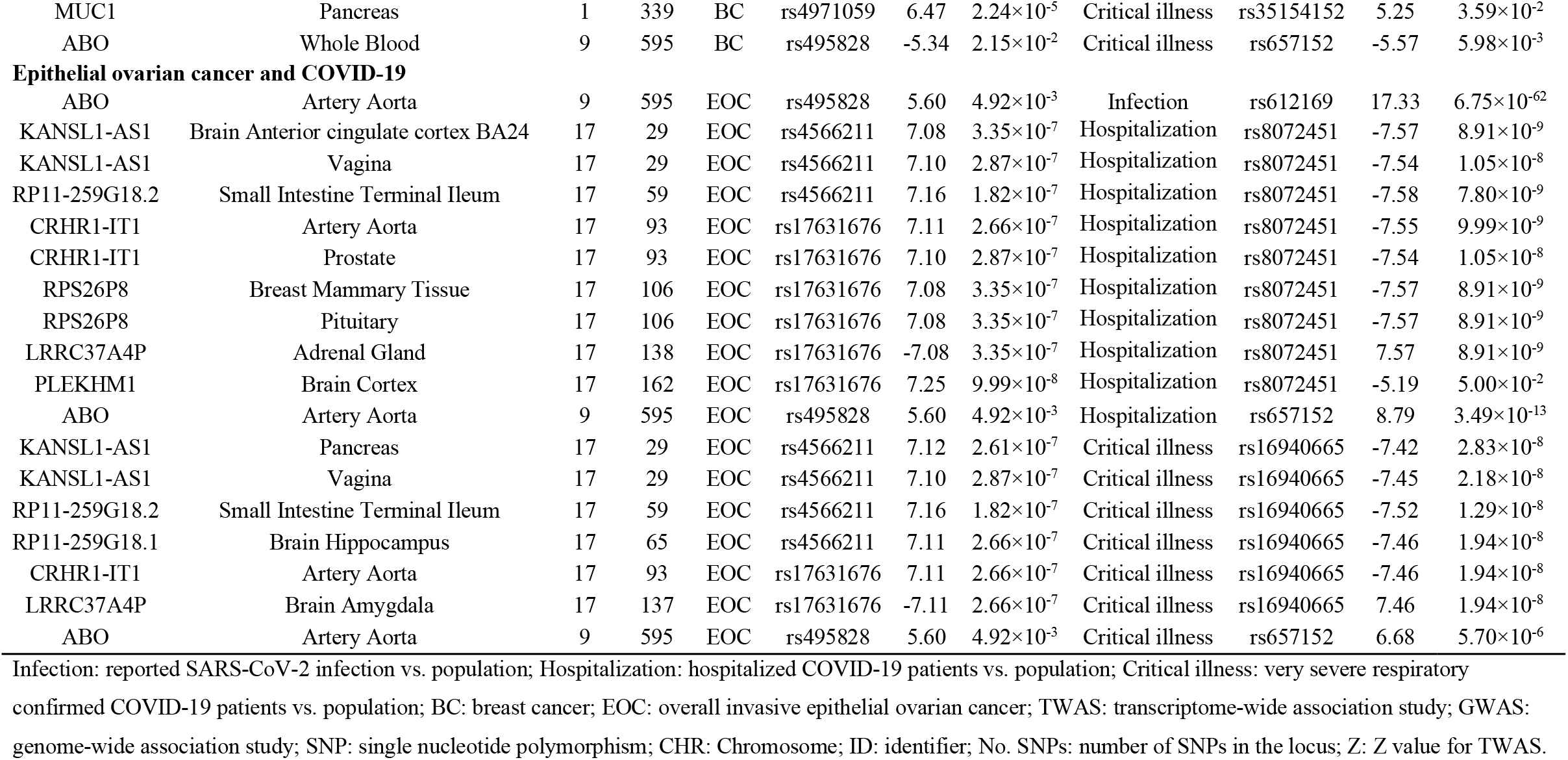
TWAS-identified shared gene-tissue pairs between COVID-19 and female-specific cancers after conditional and joint analysis.

A total of 11 genes were TWAS-significant for BC with at least one COVID-19 phenotype, including 2 with infection, 7 with hospitalization, and 7 with critical illness, enriched in tissues of adrenal gland, artery, brain, heart, pancreas, skin, stomach and whole blood. Two genes were located at pleiotropic loci identified in cross-trait meta-analysis, including *ABO* (enriched in whole blood and shared by BC with all three COVID-19 phenotypes) and *MSTO2P* (enriched in muscle skeletal and pancreas).

A total of 8 genes were TWAS-significant for EOC with at least one COVID-19 phenotype, including one with infection, 7 with hospitalization, and 6 with critical illness, enriched in tissues of artery, adrenal gland, brain, breast mammary, pancreas, and vagina. Among these TWAS significant genes, *ABO* (enriched in artery aorta and shared by EOC with all three COVID-19 phenotypes), *CRHR1-IT1* (enriched in artery aorta and prostate), and *PLEKHM1* (enriched in brain cortex) were located at pleiotropic loci identified in cross-trait meta-analysis.

### Sensitivity analysis for sample overlap

A significant genetic correlation between COVID-19 severity and EC as well as a suggestive effect of genetically predicted COVID-19 severity on EC risk were identified in the main analysis. Given the sample overlap (both GWASs contained UKB individuals), we additionally conducted a sensitivity analysis applying COVID-19 GWAS excluding UKB subjects. Results of the sensitivity analysis remained consistent with the main analysis, including directionally consistent genome-wide genetic correlation (infection: *r*_*g*_ = 0.04, *P* = 0.59; hospitalization: *r*_*g*_ = 0.14, *P* = 0.03; critical illness: *r*_*g*_ = 0.22, *P* = 1.70×10^−3^), marginal associations between genetically predicted COVID-19 phenotypes with EC risk (infection: OR_IVW_ = 1.33, 95%CI = 1.06-1.66; hospitalization: OR_IVW_ = 1.11, 95%CI = 1.03-1.1; critical illness: OR_IVW_ = 1.06, 95%CI = 1.00-1.12), as well as 2 replicated pleiotropic SNPs (SNP rs1123573 and SNP rs550057, **Supplementary Tables 15-18)**.

## Discussion

Leveraging the hitherto largest genetic data and novel statistical approaches, the current study performed a comprehensive genome-wide cross-trait analysis to systematically investigate the shared genetic influences underpinning COVID-19 and female-specific malignancies. Our study covered both the susceptibility and severity of COVID-19, as well as the three most common female cancers, BC, EOC, and EC. From a genetic perspective, our work demonstrated biological links underlying these complex traits, highlighting shared mechanisms rather than potential causal associations.

COVID-19 presents a broad spectrum of clinical manifestations ranging from asymptomatic infection to death, with the host genetic determinants one of the main influential factors^2,38^. **For COVID-19 susceptibility**, our study suggested no apparent genetic association with any of the examined female-specific cancers. **While for COVID-19 severity**, we found a significant genome-wide genetic correlation for EC with both COVID-19 hospitalization and critical illness, highlighting a non-trivial genetic component that is shared by cancer and a worse symptom of COVID-19. Notably, as the severity of infection develops, the overall COVID-19-EC genetic correlation increases, even with a decreasing sample size of corresponding COVID-19 GWAS. This may be explained by a higher level of plasma cytokines and immune responses in severe COVID-19 patients, both of which are well-established hallmarks for cancer initiation^39,40^.

A shared genetic basis can be the result of vertical pleiotropy and/or horizontal pleiotropy. In our downstream analysis performed to explore these alternatives, we identified no association of genetically predicted **COVID-19 susceptibility** with any cancer of interest, largely in line with the overall null genetic correlation. Two previous studies reported suggestive associations between genetically predicted SARS-CoV-2 infection with EC risk (OR = 1.37, 95%CI = 1.11-1.69; OR = 1.17, 95%CI = 1.01-1.34)^41,42^. However, these studies applied a limited number of IVs generated from an older version of COVID-19 GWAS (Release 4), which might reduce the precision of MR estimates due to insufficient power. With an eight-times augmented sample size of COVID-19 cases (122,616 vs. 14,134) and a larger number of IVs (16 vs. 3 or 13), our

MR did not support for such potential associations. For **COVID-19 severity**, despite suggestive associations identified for genetically predicted hospitalization or critical illness on EC risk, none withstood multiple testing. While these findings were consistent with a previous MR reporting marginal associations (hospitalization: OR = 1.15, 95%CI = 1.00-1.31; critical illness: OR = 1.08, 95%CI = 1.01-1.15)^42^, both the significance and the magnitude of estimates in our study attenuated with a nearly four-times augmented sample size of severe cases (hospitalization: 32,519 vs. 6,404; critical illness: 13,769 vs. 4,336) and a five-times enlarged number of IVs (hospitalization: 38 vs. 7; critical illness: 37 vs. 7). Based on our current findings, a worse symptom of COVID-19 does not seem to represent a risk factor for EC development, while future investigations are warranted to further establish or rule out our findings. By applying a reverse directional MR design, we further confirmed that genetically predicted female-specific cancers appear not to affect either the susceptibility or the severity of COVID-19, concordant with a previous MR study^43^.

Taken together, our MR analysis delivers timely messages that may have important clinical and public health implications. We provide evidence suggesting that COVID-19 is not likely to pose a direct effect on the immediate risk of the examined female-specific cancers, suggesting that extra cancer screening in those recovered from COVID-19 may not confer a substantial public health benefit. In fact, the potential impact of COVID-19 pandemic on routine cancer screening should be given attention, which may lead to an increased burden of cancer mortality^44^. Regarding the inconclusive effect of genetically predicted COVID-19 severity on EC risk, the marginal effect size reflects limited clinical significance. Our study, however, has not ruled out the possibility of subsequent increased risks of other chronic diseases, which, like cancer, is crucial for reducing disease burden and promoting health equity in post-COVID era^4,5^. To identify other potential long-term sequelae of COVID-19, cancers in other tissues or of other sites, cardiovascular, hematological, neurological diseases, as well as possible long-term chronic inflammation, also require attention.

Contrary to the limited genetic evidence observed for vertical pleiotropy, our cross-trait meta-analysis revealed multiple horizontal pleiotropic loci shared between cancers and COVID-19, suggesting that the previously reported phenotypic links could be largely explained by common biological mechanisms. The shared signals identified for both the susceptibility and the severity of COVID-19 further validate the notion that overall genetic correlation may fail to detect pleiotropic effects at individual variant level. Notably, many of our identified cross-trait effects were previously implicated in hematologic systems (*ABO, THBS3*)^38,45^, immune response (*WNT3, PLEKHM1, BCL11A, GON4L*)^12^, cell proliferation (*PMF1, TTC28, KANSL1*), and hormone secretion (*CRHR1, ESR1, CYP19A1*)^13,14^, reflecting potential mechanistic pathways linking COVID-19 to tumorigenesis. Via colocalization analysis, multiple genes (*ABO, WNT3, CUX2, SURF4, LCN1P2, CTD-2612A24*.*1, RP11-430N14*.*4*) showed strong evidence of a shared causal mechanism (PPH4 > 0.5). Here we highlight two interesting examples, ***ABO*** and ***WNT*3**, both are shared by COVID-19 with more than one investigated cancer.

***ABO***, a protein-coding gene involved in blood group systems biosynthesis and coagulation, is a well-known COVID-19 risk gene^38,46^. In COVID-19 patients, *ABO* contributes to hypercoagulation states and thromboses by affecting plasma glycoproteins^46-49^, meanwhile, such hypercoagulation states also frequently occurs in many cancer patients^50,51^. By regulating the circulating levels of several pro-inflammatory and immune adhesion molecules, *ABO* might contribute to both tumorigenesis and COVID-19 development^52,53^. ***WNT3*** represents a typical immune-related gene, and was identified as a shared gene for COVID-19 severity (rather than susceptibility) with cancer. By activating the WNT/β-catenin pathway, *WNT3* plays a shaping role in tumor proliferation, migration and invasion, and functions in a variety of pathological processes including inflammation, metabolism, neurological development, and fibrosis processes^54,55^. Demonstrated by previous studies, upregulation of the canonical WNT/β-catenin pathway in COVID-19 patients is associated with inflammation and cytokine storm^56^, and such inflammatory immune responses are more likely to occur in patients with severe COVID-19^39,40^. This may explain why immune-related genes such as *WNT3* were mainly identified to be shared with COVID-19 severity. Our findings suggest critical roles of coagulation and immune responses in both COVID-19 and female-specific cancers regulations, which help pinpoint therapeutic targets for both diseases.

Integrating GWAS and GTEx tissue-specific expression data, our TWAS analysis further revealed biological pleiotropy at a gene-tissue pair level. Similar with findings from CPASSOC, we found shared genes between COVID-19 and cancers that are related to hematologic systems (*ABO*), immune function (*MUC1, PLEKHM1*), and cell proliferation (*KANSL1*-*AS1*). The multiple genes identified in blood vessel or heart tissues indicate a biological mechanism through the cardiovascular system, which corroborates well with the established knowledge as both COVID-19 and cancer are associated with a number of cardiovascular complications^49,57^. In addition, we identified shared regulatory features in the nervous system, especially for COVID-19 severity. In fact, the neuro-invasiveness and neuro-invasion of SARS-CoV-2 have been well-characterized by previous studies, with more than 80% of severe COVID-19 patients showing neurological manifestations during the acute stage of their disease^58^. Through peripheral nerves and/or the hematogenous route, viruses can access the cranial nerves and influence disease manifestation^58^. Moreover, the importance of nervous system in cancer development has also been increasingly recognized^59^. Cancer cells transduce neurotransmitter-mediated intracellular signaling pathways which may lead to their activation, growth, and metastasis^59^. To sum up, these shared biologic pathways for COVID-19 and female-specific cancers implicate therapeutic strategies in clinical practice of the coexisting groups. More studies are needed to fully disclose the complex mechanisms.

Several limitations need to be acknowledged. **First**, due to unavailability of data, we conduct our analysis using sex-combined GWAS summary data of COVID-19 which may introduce sex heterogeneity. Future investigations leveraging large-scale sex-specific data may reduce this bias. **Second**, to avoid bias from population stratification, we chose GWAS data restricted to the European ancestry, limiting the generalizability to other ethnic groups. **Third**, the power of our MR analyses could still be limited by sample size, case proportion, and heritability of IVs, leading to the overall negative findings. However, by using data from the hitherto largest GWAS for COVID-19, our overall statistical power was considerably raised compared with previous genetic studies. We had 80% power at an alpha level of 0.05 to detect a 33% increased cancer risk with infection, a 43% increased risk with hospitalization, and a 47% increased risk with critical illness^60,61^. Larger GWAS data are needed to validate our results in the future. **Finally**, the delineation of COVID-19 susceptibility and severity phenotypes across studies may have been influenced by local social and medical conditions, introducing heterogeneity in the original meta-GWAS that could not be explained in our analysis.

## Conclusion

Overall, our genetic analysis extends previous findings by highlighting an intrinsic link underlying female-specific cancers and COVID-19. COVID-19 is not likely to elevate the immediate risk of female-specific cancers (BC, EOC, EC), but rather appears to share mechanistic pathways with these conditions. Such common biological mechanisms are specifically substantiated by the pleiotropic loci and shared genes identified in our study, implicating therapeutic strategies for future clinical practice.

## Data Availability

This study did not generate new datasets or codes. All data used in our study are publicly-available. GWAS summary statistics of the COVID-19 Host Genetics Initiative are accessible at https://www.covid19hg.org/. GWAS summary statistics for breast cancer, epithelial ovarian cancer, and endometrial cancer can be downloaded from the GWAS catalog (https://www.ebi.ac.uk/gwas/) or from the websites of the consortium (http://bcac.ccge.medschl.cam.ac.uk/bcacdata/, http://ocac.ccge.medschl.cam.ac.uk/). More details of the approaches as well as the codes are available at https://github.com/bulik/ldsc (LDSC), https://mrcieu.github.io/TwoSampleMR/ (TwoSampleMR), http://hal.case.edu/~xxz10/zhuweb/ (CPASSOC), https://github.com/hailianghuang/FM-summary (FM-summary), https://chr1swallace.github.io/coloc/ (Coloc), https://www.cog-genomics.org/plink/1.9/ (PLINK), https://grch37.ensembl.org/info/docs/tools/vep/index.html (VEP), http://gusevlab.org/projects/fusion/ (FUSION).

## Acknowledgments

We thank all the patients, staff and investigators who contributed to the COVID-19 Host Genetics Initiative, BCAC consortium, OCAC consortium and ECAC consortium. This study was supported by funds from the National Natural Science Foundation of China (81874283, 81673255, 81874282), the National Key R&D Program of China (2020YFC2006505), the Health Commission of Sichuan Province (20PJ093), the Key R&D Program of Sichuan, China (2022YFS0055), the Recruitment Program for Young Professionals of China, the Promotion Plan for Basic Medical Sciences, the Development Plan for Cutting-Edge Disciplines, Sichuan University, and other Projects from West China School of Public Health and West China Fourth Hospital, Sichuan University.

## Data availability

This study did not generate new datasets or codes. All data used in our study are publicly-available. GWAS summary statistics of the COVID-19 Host Genetics Initiative are accessible at https://www.covid19hg.org/. GWAS summary statistics for breast cancer, epithelial ovarian cancer, and endometrial cancer can be downloaded from the GWAS catalog (https://www.ebi.ac.uk/gwas/) or from the websites of the consortium (http://bcac.ccge.medschl.cam.ac.uk/bcacdata/, http://ocac.ccge.medschl.cam.ac.uk/). More details of the approaches as well as the codes are available at https://github.com/bulik/ldsc (LDSC), https://mrcieu.github.io/TwoSampleMR/ (TwoSampleMR), http://hal.case.edu/∼xxz10/zhuweb/ (CPASSOC), https://github.com/hailianghuang/FM-summary (FM-summary), https://chr1swallace.github.io/coloc/ (Coloc), https://www.cog-genomics.org/plink/1.9/ (PLINK), https://grch37.ensembl.org/info/docs/tools/vep/index.html (VEP), http://gusevlab.org/projects/fusion/ (FUSION).

